# A workflow for clinical profiling of BRCA genes in Chilean breast cancer patients via targeted sequencing

**DOI:** 10.1101/2024.09.25.24314295

**Authors:** Evelin González, Rodrigo Moreno Salinas, Manuel Muñoz, Soledad Lantadilla Herrera, Mylene Cabrera Morales, Pastor Jullian, Waleska Ebner Durrels, Gonzalo Vigueras Stari, Javier Anabalón Ramos, Juan Francisco Miquel, Lilian Jara, Carol Moraga, Alex Di Genova

## Abstract

**Background:** Breast cancer (BC) is the leading cause of cancer-related deaths among women globally and in Chile. Mutations in the tumor-suppressor genes BRCA1 and BRCA2 significantly increase the risk of developing cancer, with the probability rising by more than 50%. Identifying pathogenic variants in BRCA1 and BRCA2 is crucial for both diagnosis and treatment. Targeted panels, which focus on medically relevant subsets of genes, have become essential tools in precision oncology. Beyond technical and human resource factors, standardized bioinformatics workflows are essential for the accurate interpretation of results. We developed a robust bioinformatics pipeline, implemented with Nextflow, to process sequencing data from targeted panels to identify germline variants.

**Results:** We developed an automated and reproducible pipeline using Nextflow for the targeted sequencing of BRCA1/2 genes. The pipeline incorporates two variant callers, Strelka and DeepVariant, both of which have demonstrated high performance in detecting germline SNVs and indels. The runtime is efficient, with a median execution time of less than 3 minutes per task. We sequenced and processed 16 samples from breast cancer patients. In our analysis, we identified 8 nonsynonymous mutations in *BRCA1* and 9 in *BRCA2*. Of the total reported germline mutations, 97% were classified as benign, 1% as pathogenic, 1% as of uncertain significance, and 1% as unknown. The allelic frequencies observed in our cohort closely resemble those of Admixed American and South Asian populations, with the greatest divergence observed in comparison to African individuals.

**Conclusion:** We successfully analyzed the *BRCA1* and *BRCA2* genes in 16 breast cancer patients at a public hospital in Chile. A custom Nextflow pipeline was developed to process the sequencing data and evaluate the pathological significance of the identified genetic variants. By employing multiple variant-calling methodologies, we were able to detect and mitigate potential false positives, thereby enhancing the accuracy and reliability of variant detection through cross-verification. A pathogenic variant was identified in one patient, while benign or likely benign variants were found in the remaining 15. Expanding the number of oncogenes sequenced per patient could improve the detection of actionable variants.

## Background

Breast cancer is a significant public health issue and the most common oncological pathology among women (Bray et al. 2024). In Chile the incidence of breast cancer was 210 cases per 100,000 women, higher than the reported by GLOBOCAN for 2020 (Villalobos et al. 2023). In 2023, a total of 7,503 women insured by the public health system were confirmed to have breast cancer, with 77% of them being over 50 years old (Merino-Pereira 2023).

Breast cancer is characterized as a heterogeneous group of tumors that can be classified based on histopathological features, genetic alterations, and gene-expression profiles (Kreike et al. 2007). These tumors develop from aberrant gene expression caused by mutations in the DNA. While environmental and lifestyle factors play a crucial role in DNA mutations, genetic factors can also increase the risk of developing cancer. For example, mutations in the breast cancer genes BRCA1 and BRCA2 raise the risks for breast, ovarian, fallopian tube, and peritoneal cancer (Acevedo et al. 2023).

Although the incidence rate varies by ethnicity and geographic regions (Fackenthal and Olopade 2007), between 10 to 50% of patients with hereditary breast cancer carry deleterious mutations at *BRCA1* or *BRCA2,* which increase their risk of developing breast cancer in more than 50% (Antoniou et al. 2003; Chen and Parmigiani 2007; Kuchenbaecker et al. 2017). In Chile, it has been estimated that 15 to 20% of patients with hereditary breast cancer carry deleterious mutations in *BRCA1* or *BRCA2 (Alvarez et al. 2017; Gallardo et al. 2006; Jara et al. 2006)*. Identifying pathogenic variants in BRCA1 and BRCA2 is crucial for determining genetic predisposition and assessing the risk of potentially developing breast cancer. Furthermore, identifying pathogenic variants is essential for prevention strategies, treatment planning, and the development of therapeutic drugs (Lee, Moon, and Kim 2020; Tung et al. 2020). For example, in 2018 the Food and Drug Administration (FDA) approved the use of olaparib for the treatment of patients with HR-positive/HER2-negative metastatic breast cancer who have a germline BRCA mutation (Schwartzberg and Kiedrowski 2021). Targeted panels that sequence medically relevant subsets of genes have become a fundamental component of precision oncology (Koboldt 2020).

In Chile, there have been advances in precision oncology focused on BRCA1/2 genes. In 2006, the first genetic study of BRCA gene mutations was published (Jara et al. 2006). Among high-risk breast and/or ovarian cancer families, 10.9% were found to carry BRCA1 mutations, while 4.7% carried BRCA2 germline mutations (Jara et al. 2006). A 2017 study screened 453 Chilean patients with hereditary breast cancer for mutations in BRCA1/2, finding a total of 25 mutations (6 novel) in 71 index patients, with nine mutations present exclusively in Chilean patients (Alvarez et al. 2017). The Fundación Arturo López Pérez (FALP) conducted the latest NGS sequencing study on breast cancer in Chile(Martin et al. 2024). A multigene panel profiling on 722 patients with breast or ovarian cancer identified pathogenic variants in the BRCA1/2 genes in 103 (14%) individuals, including seven previously unreported BRCA1 variants (Martin et al. 2024).

Current breast cancer (BC) guidelines in Chile lack defined strategies for germline genetic testing. A 2023 study found that only 15% of BC patients who meet NCCN criteria for germline testing actually receive it (Acevedo et al., 2023). This percentage is even lower in public hospitals. Extrapolating these figures to the entire country suggests that fewer than 1 in 10 individuals who meet the NCCN criteria have access to testing within the public health system (Acevedo et al., 2023).

Technological advances have made genetic sequencing increasingly accessible at a lower cost. In Chile, certain institutions have the capacity and expertise to implement NGS protocols within their facilities. These capabilities can serve as a foundation for integrating these technologies into genetic diagnostics (Marcelain et al. 2019). In addition to the need for equipment, reproducible bioinformatics methodologies and workflows are essential. These should encompass data processing, quality control, variant calling, annotation, visualization, and ultimately reporting in a format that is readily interpretable for clinical decision-making and genetic counseling (Jäger 2022). Accurate variant calling in NGS data is a critical step upon which virtually all downstream analysis and interpretation processes depend (Koboldt 2020). Additionally, employing standardized workflows that are publicly available to the community could reduce costs. Here, we performed targeted sequencing of the BRCA1/2 genes in 16 breast cancer patients at a public hospital in Chile. Utilizing a custom-designed Nextflow pipeline, we processed the sequencing data to identify, annotate, and assess the impact of genetic variants in the BRCA1/2 genes. Our findings are intended to help medical practitioners guide more targeted interventions for at-risk breast cancer patients and demonstrate that NGS technologies can be effectively implemented in public hospitals in Chile.

## RESULTS

### 1. A Nextflow Workflow for BRCA1/2 variant profiling

An NGS workflow for paired-end Illumina libraries was developed to identify germline variants for targeted sequencing using the AmpliSeq for Illumina BRCA Panel (Tariq et al. 2021). This workflow, built on the Nextflow platform, enables an autonomous, reproducible, and scalable scientific pipeline (Patel et al. 2024). The integrated tools are executed through containers, and the entire workflow is available on the GitHub repository ( https://github.com/digenoma-lab/BRCA).

The BRCA workflow (Figure 1) starts by mapping FASTQ files to the human hg38 genome reference using the Burrows-Wheeler Alignment tool (BWA) (H. Li and Durbin 2009) and performs read quality control with FastQC, which allows for the identification of potential problems in the generated reads. Metrics of aligned reads and exon coverage are computed with Qualimap (García-Alcalde et al. 2012). After aligning reads with BWA during data preprocessing, the reads are processed to ensure consistency and correct mate information in BAM files using Samtools fixmate, and reads are sorted using the Samtools sort (Danecek et al. 2021). In parallel to variant calling, the outputs from FastQC, Qualimap, and Samtools are consolidated by MultiQC (Ewels et al. 2016) to create an HTML report with the metrics of processed samples. The preprocessed BAM files are used as input for Variant calling that is performed using two callers: Strelka (Kim et al. 2018) and DeepVariant (Poplin et al. 2018; Yun et al. 2021). Both have demonstrated high performance for detecting germline SNVs and Indels. These callers were executed in two modes: single-sample mode (S), which calls variants for each sample individually, and multisample mode (MS), which performs joint variant calling and genotyping, considering all samples simultaneously. Finally, the functional annotation of the identified variants is performed using ANNOVAR (Wang, Li, and Hakonarson 2010), integrating databases such as gnomAD v4, dbSNP v150, ICGC v28, ClinVar, InterVar, and REVEL (Figure 1). REVEL is an ensemble method for predicting the pathogenicity of missense variants based on 18 individual tools, including conservation and functional scores (Ioannidis et al. 2016) and InterVar is a tool designed for the clinical interpretation of genetic variants according to the ACMG/AMP 2015 guidelines (Q. Li and Wang 2017). We used REVEL and InterVar to understand the functional significance of the genetic variants and their potential effects on BRCA12 genes. This pipeline design ensures comprehensive variant detection and annotation, providing a robust and reliable tool for clinical and research applications.

**Figure 1.**
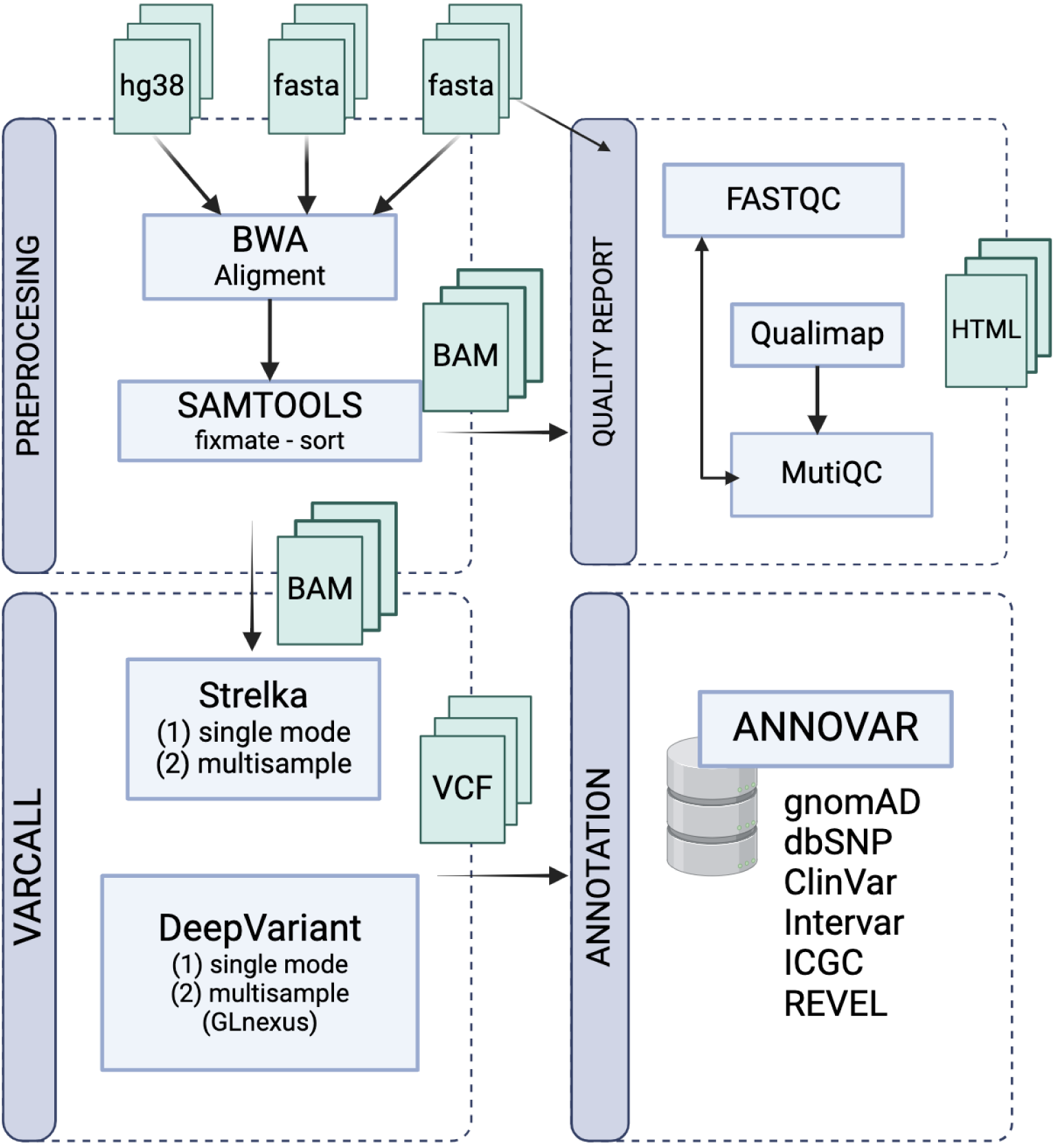
Nextflow pipeline for detecting SNVs and Indels in BRCA1/2 genes. The workflow identifies germline variants from paired-end sequencing data and includes four main processes: preprocessing, quality reporting, variant calling, and annotation. Subprocesses are shown in rectangles, with input and output files in green.

### 2. The BRCA pipeline enables rapid and reproducible variant profiling for clinical practice

Sixteen breast cancer patients were sequenced using the AmpliSeq for Illumina BRCA Panel, which includes all exonic regions of the *BRCA1* and *BRCA2* genes and flanking intronic sequences, covering a total of 22 Kb (Tariq et al. 2021).

To ensure reproducibility, we executed the BRCA Nextflow pipeline 10 times using data from 16 breast cancer patients. The results showed high reproducibility, with 100% concordance of the variants reported by Strelka and DeepVariant in both single-sample and multi-sample modes. Additionally, we used the Fastq Shuffle tool (Sanders 1998) to create new FASTA files by randomly reordering the reads per sample for three independent runs, as well as altering the sample names in one instance. We observed a 100% concordance of the variants reported in Strelka and DeepVariant in both modes. Finally, all tasks showed a median execution time of less than 3 minutes (Supplementary Figure 1) and with an average total execution of X minutes for all 16 samples. The most time-consuming tasks were annotation with ANNOVAR, which had a median of 2 minutes and 43 seconds, and BWA-MEM, with a median of 1 minute and 31 seconds. Among the variant callers, Strelka had a median execution time of 50 seconds in single-sample mode and 1 minute and 35 seconds in multi-sample mode. In comparison, DeepVariant had a median execution time of 51 seconds, and GLnexus took only 4.5 seconds per run (Supplementary Figure 1). Overall, the BRCA Nextflow pipeline demonstrated exceptional performance, with rapid execution times and high reproducibility, making it a reliable and efficient tool for clinical practice.

### 3. High-quality sequencing and comprehensive variant report for BRCA1/2 *genes*

We obtained high-quality reads for all 16 samples, with an average of 478,730 raw reads per sample (Supplementary Figure 2). All the reads passed quality control. The average GC content was 38%, with 99% of reads mapped to the target regions per sample, achieving a mean coverage of 1,905X (Supplementary Figure 2). We identified a total of 70 unique mutations in the BRCA1/2 genes across all patients, of which 4 are UTR, 39 intronic, and 27 exonic. Among the exonic mutations, 10 are silent, 16 are missense, and one is a frameshift insertion Figure 2(). The most frequent mutation changes are T>C (n=193), followed by C>T (n=81) and T>G (n=26). Strelka and DeepVariant identified a median of 13 mutations in exonic regions per patient. Patients LV and PV had a higher number of mutations (n=9), while patients RQ had 3 mutations and DC had 2 mutations. (Figure 2). We examined coverage variation across the target regions of the panel and observed a high degree of uniformity (Supplementary Figure 3).

**Figure 2:**
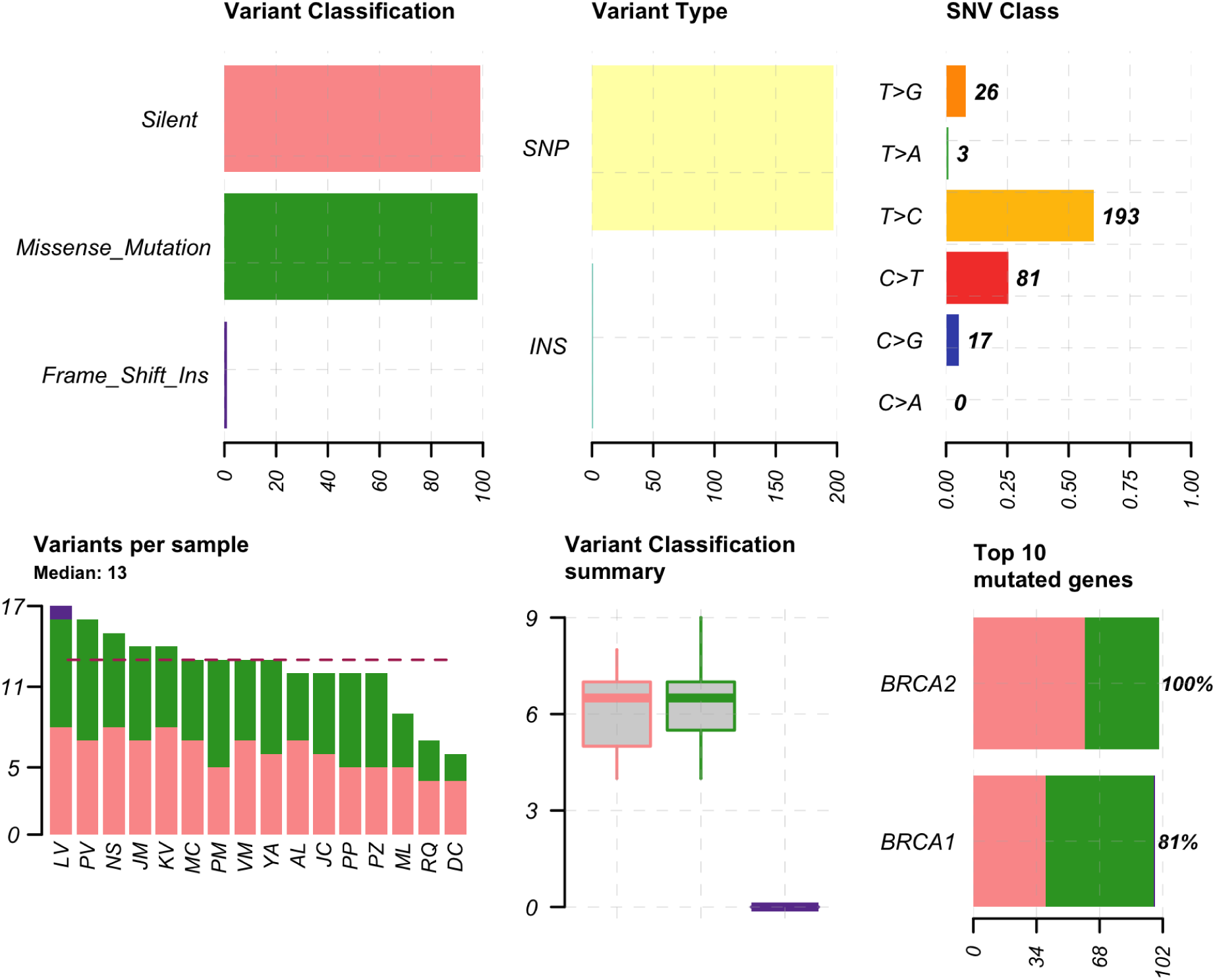
Summary of germline mutations found in BC patients. These panels present the classification, type, and distribution of variants in 16 breast cancer patients, including Silent, Missense Mutations, and Frame Shift Insertions. Details SNV base changes, and shows a median of 13 variants per sample. BRCA2 and BRCA1 have mutation rates of 100% and 81%, respectively.

**Figure 3:**
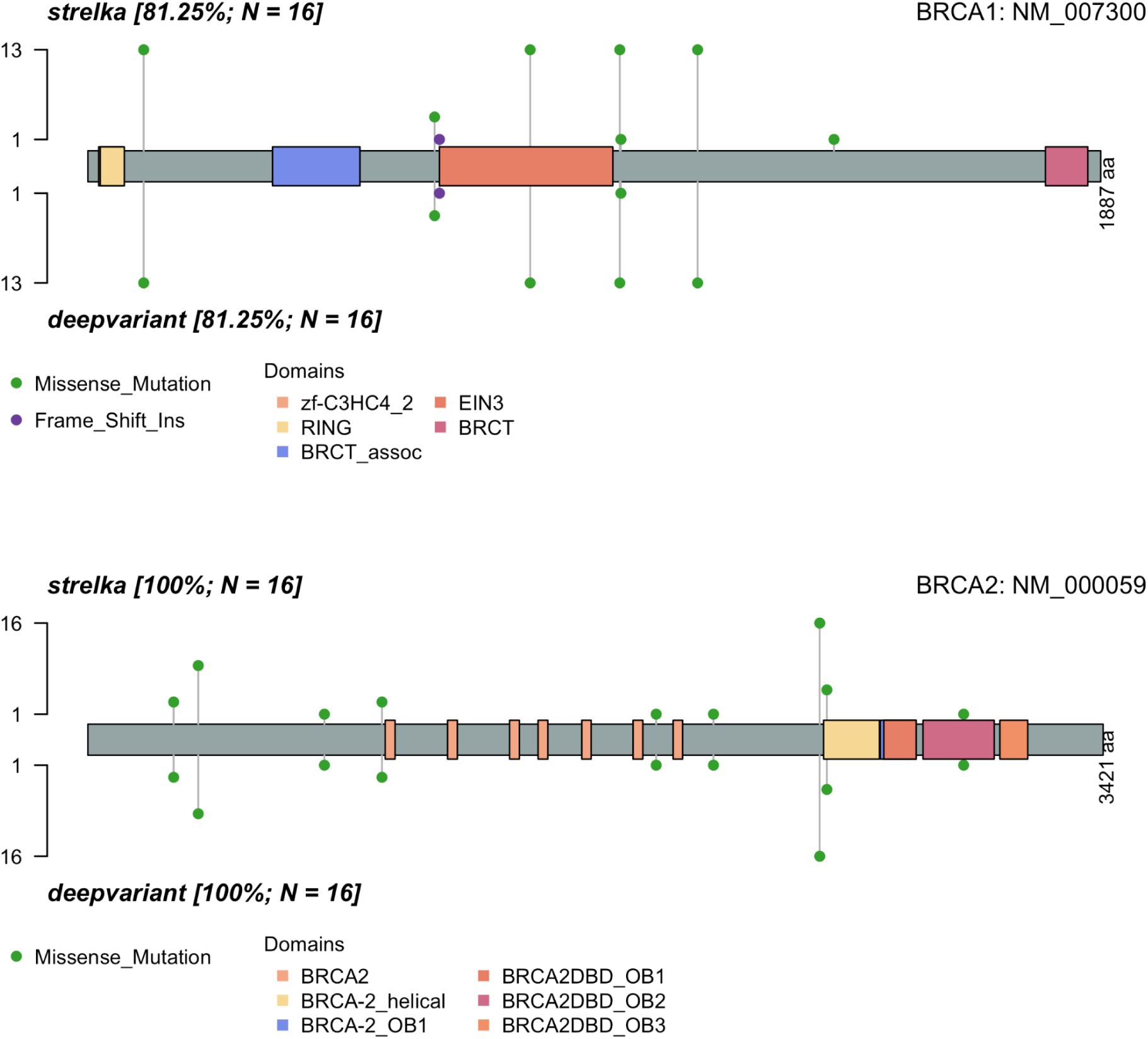
Lollipop Plots of variants in *BRCA1* and *BRCA2* detected by Strelka and DeepVariant callers. Graphical representation of the *BRCA1* and *BRCA2* genes, comparing the mutations obtained by Strelka and Deepvariant. The Y-axis shows the number of patients with missense mutations (green) and frameshift insertions (purple). The top section represents the results from Strelka, and the bottom section represents the results from DeepVariant of each gene. The domains are in the labels. The results correspond to the variants obtained by both variant callers in single mode.

### 4. Comparison of Single and Multi-Sample Modes in Variant Detection for BRCA1 and BRCA2 Using Strelka and DeepVariant

In Strelka, both single and multi-sample modes found 8 nonsynonymous variants in BRCA1. These were 1 frameshift insertion and 7 missense mutations. Moreover, 9 missense mutations in *BRCA2 are also found by both strelka modes*. The E1390G variant in *BRCA1* was identified in one patient in single mode and in three patients in multisample mode. This suggests it could be a novel mutation or a sequencing artifact (Supplementary Figure 4). Visual inspection of this variant in IGV supports the former rather than the latter possibility (variant phred quality 49, Supplementary Figure 6).

On the other hand, DeepVariant, in both single and multi-sample modes, reported identical exonic nonsynonymous variants. We observed no differences in variant calling between the two modes of DeepVariant (Supplementary Figure 5).

### 5. Frequency and Impact of BRCA1/2 Mutations in Chilean Patients Diagnosed with Breast Cancer

The 16 sequenced patients were Chilean women, up to 40 years old, diagnosed with breast cancer via histopathological confirmation between January 2015 and December 2021. None of the participants were selected based on a family history of cancer. Of the germline mutations detected on this cohort, 97% are benign, 1% are pathogenic, 1% are of uncertain significance, and 1% are unknown. We identified a pathogenic variant in one breast cancer patient (LV). Both Strelka and DeepVariant reported the p.L655Ffs*10 frameshift insertion in exon 9 of BRCA1 in single and multisample modes (Figure 2 and 4). This variant, with rs80357880, is classified as pathogenic in ClinVar and has been associated with hereditary breast and ovarian cancer. According to the OncoKB database (Chakravarty et al. 2017), this variant is classified as likely oncogenic with a Level 1 evidence classification, indicating that it is an FDA-recognized biomarker predictive of response to several FDA-approved drugs, including PARP inhibitors.

**Figure 4:**
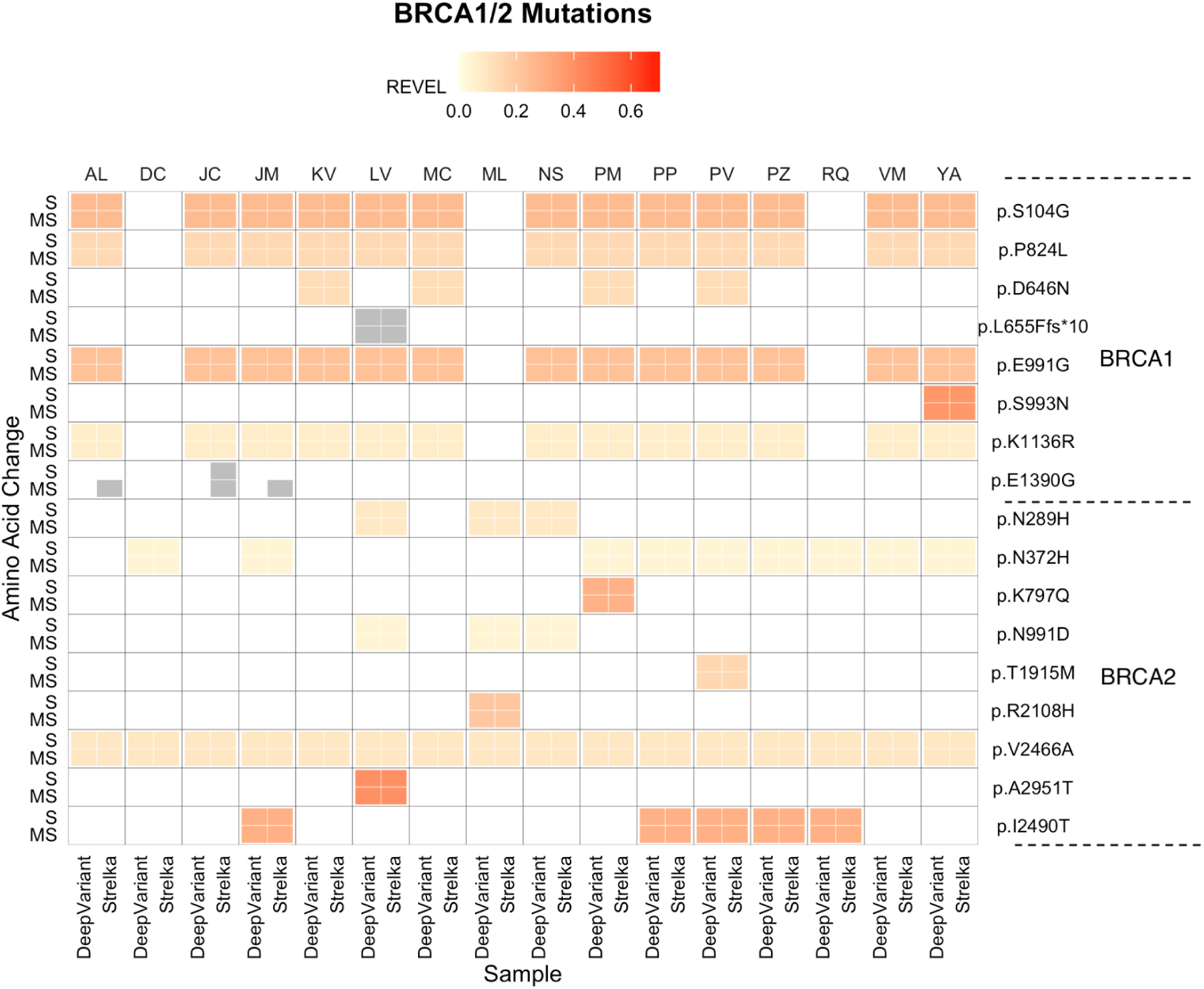
Heatmap of BRCA1/2 mutations reported in 16 breast cancer patients using DeepVariant and Strelka, both in single and multisample mode. The Y-axis shows the amino acid changes (right) in single and pooled modes (left), and the X-axis shows the 16 patients (top) across the two varcallers used (bottom). The color indicates the potential pathogenicity as given by the REVEL score. Variants without REVEL score are in gray. Single-sample mode (S), Multisample mode (MS).

In patient PM, we identified a *BRCA2* mutation of uncertain significance, p.K797Q, reported by both Strelka and DeepVariant in both modes. According to ClinVar, this mutation has been associated with hereditary breast and ovarian cancer syndrome. Predictors SIFT, PROVEAN, and M-CAP suggest the variant could be deleterious. However, FATHMM, REVEL (score of 0.281), and InterVar indicate that it could be benign. Additionally, this mutation is not present in gnomAD v4.0. We identified one variant that has not been detected in other repositories. The E1390G mutation in *BRCA1* was reported by Strelka in single mode for one patient and in multisample mode for three patients. DeepVariant did not report this variant. This mutation does not have a REVEL score; however, predictors MutationTaster, MetaRNN, and FATHMM-MKL classify it as benign. The E1390G mutation in BRCA1 was visually inspected using IGV for the three patients reporting this variant. The mutation is located in reads flanking one of the target regions, showing low read coverage (greater than 8). However, it has a Strelka Phred quality score of 47, suggesting a low probability of being an artifact (Supplementary Figure 6). The mutations with the highest REVEL scores are A2951T in *BRCA2* and S993N in *BRCA1*, with scores of 0.39 and 0.37, respectively. We did not identify any variants with a REVEL score over 0.5 (Figure 4), which is the minimum required to classify variants as pathogenic.

We identified 14 mutations in BRCA1/2 that are present in gnomAD. The most frequent mutations in our cohort for *BRCA1* are K1136R, E991G, P824L, and S104G, each reported in 13 patients, with an allele frequency of 0.5 in our cohort (Figure 4). For *BRCA2*, the most common mutations are I2490T, N372H, and V2466A, found in 5, 9, and 16 patients, respectively, with allele frequencies of 0.15, 0.34, and 1.0 in our cohort (Figure 4). In the gnomAD Latino/Admixed American (AMR) subpopulation, the frequencies for these mutations are 0.07, 0.30, and 0.99, respectively.

To evaluate the frequency of these mutations, we compared the allele frequencies (AF) in our Chilean cohort (CHI) with those of the subpopulations reported in gnomAD. We subsequently performed a PCA analysis based on the allele frequencies of the mutations in our cohort and gnomAD, followed by hierarchical clustering. We observed that the first principal component accounted for 44.9% of the variance in allele frequencies, while the second explained 18%. Using the first two components (62.9% variance), we identified three distinct clusters. This analysis indicates that the allele frequencies identified in our study are more closely aligned with those reported for South Asians and Admixed Americans (clustered in blue in Figure 5). The mutations contributing most to the variance of dimension 1 were E991G, K1136R, and S104G with contributions of 12.8%, 12,6%, and 12,6%, respectively (Figure 5).

**Figure 5:**
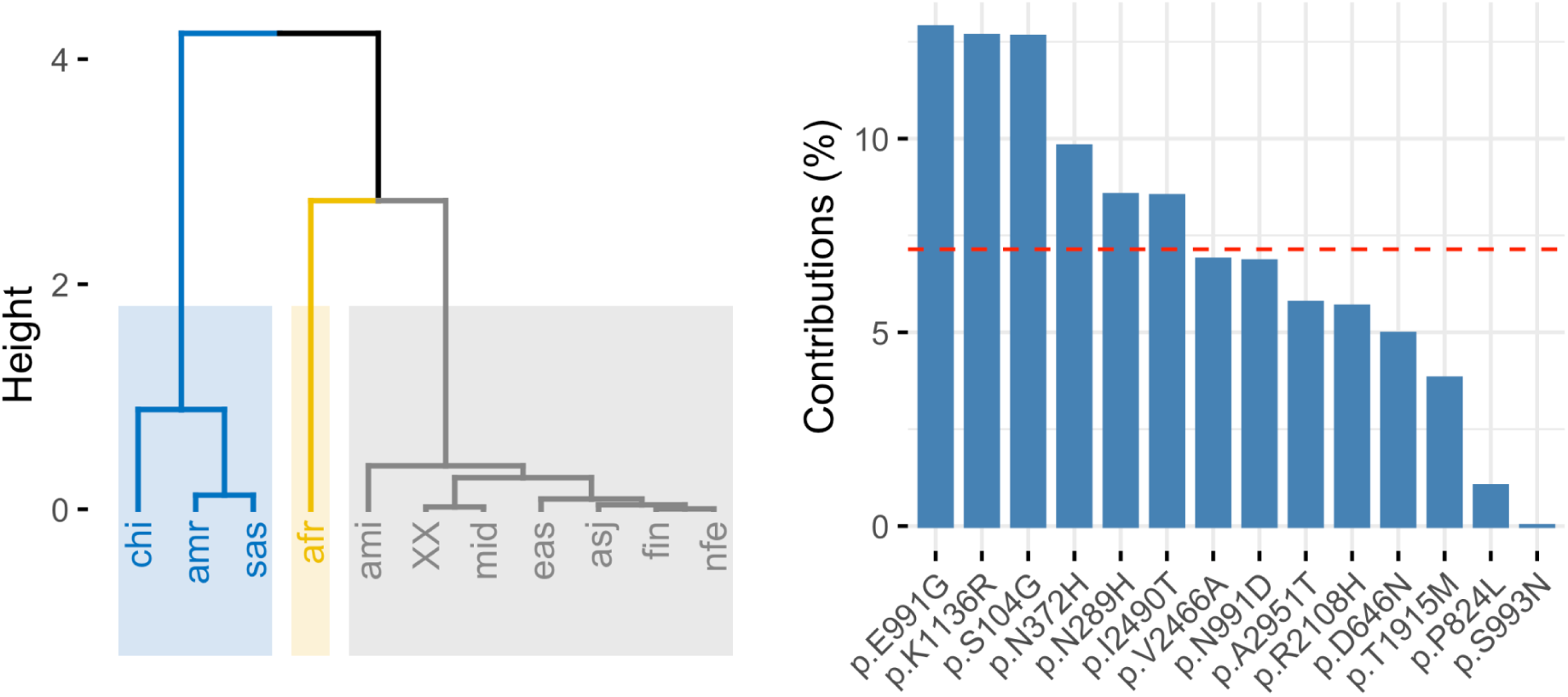
Hierarchical clustering and variants contribution on principal components. A) Clustering of PCA dimensions 1 and 2 B) Contribution of mutations to dimensions 1 and 2. A dashed line shown corresponds to the expected value if the contribution were uniform. Subpopulations: Africans (AFR), Admixed Americans (AMR), Amish (AMI), Ashkenazi Jewish (ASJ), East Asians (EAS), South Asians (SAS), Finns (FIN), Middle Easterners (MID), Non-Finnish Europeans (NFE), and women (XX).

## Discussion

Breast cancer continues to be one of the leading causes of cancer-related mortality among women worldwide, including in Chile. Genetic predisposition, particularly through mutations in the BRCA1 and BRCA2 genes, plays a pivotal role in increasing the risk of breast and other related cancers. Our study aimed to develop an automated and reproducible pipeline for identifying pathogenic variants in the BRCA1 and BRCA2 genes among Chilean breast cancer patients, with the goal of improving genetic testing at a public hospital in Chile.

Mutations in BRCA1 and BRCA2 genes are well-documented as major contributors to hereditary breast cancer, with incidence variations observed based on ethnicity and geography. In Chile, approximately 15-20% of patients with hereditary breast cancer carry deleterious mutations in these genes. Our findings are consistent with these statistics, reinforcing the importance of genetic testing and counseling within our population (Acevedo et al. 2023). Despite advancements in technology making genetic sequencing more accessible and affordable, challenges remain in integrating these tools into routine clinical practice in Chile. Consequently, fewer than 10% of individuals who meet the NCCN criteria have access to testing in the public health system. The latest study by Fundación Arturo López Pérez (FALP) highlights the potential of next-generation sequencing (NGS) for identifying pathogenic variants in breast cancer patients (Martin et al. 2024). However, broader implementation of sequencing technologies requires addressing challenges related to infrastructure, reproducible bioinformatics workflows, and clinical interpretation of genetic data (Yadav et al. 2023).

We developed a custom and robust Nextflow NGS workflow for processing the AmpliSeq Illumina BRCA Panel data. Our pipeline ensures reproducibility, demonstrated by 100% concordance in variant calling across multiple runs and perturbation experiments (read shuffling and label rename). The integration of tools such as Strelka and DeepVariant for variant calling, along with ANNOVAR for annotation, enhances the sensitivity and reliability in identifying and interpreting genetic variants. We identified a total of 70 unique mutations in BRCA1/2 genes, including novel and known pathogenic variants. Notably, the p.L655Ffs*10 frameshift insertion in *BRCA1*, classified as pathogenic, underscores the critical need for genetic testing to guide clinical decision-making. Additionally, our study identified a variant of uncertain significance, emphasizing the complexity of interpreting genetic data and the necessity for continuous updates in local genetic databases and predictive tools.

Significantly, our sequencing was performed at a public regional hospital, demonstrating for the first time that comprehensive BRCA testing is feasible in such settings. This opens the door to expanding genetic testing to whole-genome sequencing, which is particularly needed to address patients who test negative for BRCA mutations but may have other genetic predispositions to breast cancer. Our principal component analysis revealed that the allele frequencies of BRCA1/2 mutations in our cohort are closer to those reported for South Asians and Admixed Americans in gnomAD. This finding highlights the importance of considering population-specific genetic variations in developing tailored prevention and treatment strategies.

To enhance the impact of genetic testing in Chile, several measures can be employed: first, update national breast cancer guidelines to include mandatory genetic testing for high-risk individuals as our cohort (women with breast cancer before 40 of age), strengthen the technological capacity of local institutions to implement NGS protocols and bioinformatics workflows, promote training programs for healthcare professionals in genetic counseling and the interpretation of genetic data, and encourage collaborative research efforts to continuously update and validate genetic databases with Chilean specific data.

Our workflow is a step towards the use of open-source software in clinical practice, which allows for minimizing costs per patient. For variant calling, best practices in clinical sequencing recommend incorporating 2 or 3 tools for each class of variant to maximize detection sensitivity (Koboldt 2020). We implemented two high-accuracy varcallers to detect SNPs and indels. Strelka2, in the PrecisionFDA Consistency and Truth Challenge, improved the indel F-score of the best submission by 0.11% (Kim et al. 2018) and DeepVariant won the highest performance award for SNPs at the PrecisionFDA Truth Challenge 2016 (Poplin et al. 2018)

The discordance between the implemented callers allows us to identify potential artifacts that could be erroneously reported as variants. The E1390G mutation in *BRCA1* was the only discordant variant between both variant callers. A subsequent visual inspection with IGV identified it as a potential variant (Supplementary Figure 6). However, analytical validation is necessary to adhere to the standards and guidelines for the NGS bioinformatics pipeline. Improperly developed, validated, and/or monitored pipelines may generate inaccurate results that could have negative consequences for patient care (Roy et al. 2018).

In summary, our study demonstrates the feasibility and importance of integrating advanced genetic testing into clinical practice for breast cancer in Chile. By addressing existing challenges and leveraging technological advancements, we can significantly improve early detection, prevention, and personalized treatment strategies, ultimately enhancing patient outcomes.

## Material and Methods

### Patients and sample collection

We analyzed the DNA of 16 breast cancer patients from a cohort study in the O’Higgins region of Chile, treated at the Hospital Regional in Rancagua. Inclusion criteria were Chilean patients, males of all ages and females up to 40 years old, diagnosed with breast cancer with histopathological confirmation between January 2015 and December 2021. None of the participants were selected based on a family history of cancer. This study represents the first geographically based cancer study in Chile. The Comité Ético Científico del Servicio de Salud Metropolitano Sur (Ethics Committee of the South Metropolitan Health Service) reviewed and approved this study. Informed written consent was obtained from all participants, who also received pre- and post-genetic counseling in accordance with international recommendations, as well as follow-up by geneticists. Swabs samples were collected from each patient, and DNA was extracted using standard protocols.

### DNA Extraction and Sequencing

Genomic DNA was extracted using the QIAamp DNA Mini Kit (Qiagen) following the manufacturer’s instructions. The quality and quantity of the extracted DNA were evaluated using a NanoDrop 2000 spectrophotometer (Thermo Fisher Scientific) and a Qubit 3.0 Fluorometer (Invitrogen). Next-generation exome sequencing (NGS) of the BRCA1 and BRCA2 genes was performed using the Illumina iSeq 100 platform at the Molecular Laboratory of the Pathological Anatomy Service at the Hospital Regional de Rancagua, following the recommended protocols for the Illumina AmpliSeq BRCA Panel. Briefly, 10 ng of genomic DNA from each sample was used for library construction, followed by target amplification, library purification, and quantification. Sequencing was performed on iSeq100 platform, generating paired-end reads of 150 bp.

### Bioinformatics Workflow

A custom Nextflow pipeline was developed to process the sequencing data, ensuring reproducibility and scalability. The workflow comprised the following steps: 1. Quality Control: Raw sequencing reads were subjected to quality control using FastQC to identify potential issues in the data. 2. Alignment: Reads were aligned to the human reference genome (hg38) using the Burrows-Wheeler Aligner (BWA-MEM2). 3. Post-alignment Processing: Aligned reads were processed to ensure consistency and correct mate information using Samtools fixmate and sorted with Samtools sort. 4. Variant Calling: Variants were called using two tools, Strelka and DeepVariant, in both single-sample and multisample modes. GLnexus was utilized to consolidate variant calls from GVCFs DeepVariants files. 5. Mapping Quality: Metrics of aligned reads and coverage were analyzed with Qualimap, and consolidated reports were generated using MultiQC. 6. Variant Annotation: Identified variants were annotated using ANNOVAR, integrating multiple databases such as gnomAD, dbSNP, ICGC, ClinVar, and REVEL.

### Reproducibility and Performance Analysis

To ensure the reproducibility of our pipeline, we performed ten independent runs using the same data from the 16 breast cancer patients. Additionally, the Fastq Shuffle tool was used to create new FASTA files by randomly reordering reads per sample for three independent runs, with one instance involving the alteration of sample names. Execution times for each task were recorded using trace information provided by Nextflow. The bcftools isec was employed to check the reproducibility of results.

### Quality Control Metrics

To verify the quality of sequencing, we collect multiple metrics throughout the workflow using FASTQC, Qualimap, and Samtools. FASTQC reports the total number of sequencing reads, duplicated reads, and the quality scores (in Phred scale) for both forward and reverse reads. Qualimap provides metrics such as percentage of GC content, insert size, percentage of reads with at least 10x-50x coverage, mean coverage, and total reads per sample. Samtools indicates the number of mapped reads. All these metrics are summarized and visualized in an HTML report generated by MultiQC.

We evaluated the quality of sequencing reads, coverage, and variant calling performance. High-quality reads were obtained for all samples, with an average of 478,730 raw reads per sample and a mean coverage of 1,905X. Variants were classified into silent, missense, frameshift insertions, and intronic mutations. The functional significance of identified variants was assessed using REVEL and *in silico* predictors.

### Statistical Analysis

Principal component analysis (PCA) and clustering was conducted to compare allele frequencies of BRCA1/2 mutations in our cohort with those in the gnomAD database. The first two principal components were analyzed to explain the total variance, and contributions of individual mutations to these dimensions were calculated.

### Data Visualization

Data visualization was performed using various tools: Integrative Genomics Viewer (IGV) was used for visual inspection of alignments for clinically relevant variants. R Statistical analysis and plots, such as bar plots, lollipop plots, and heatmaps, were generated using R programming language. For processing variants and their functional information, annotation files were transformed into MAF format using the maftools library in R (Mayakonda et al. 2018). This library was also utilized to summarize, analyze, and visualize the data, including generating lollipop plots for *BRCA1/2* and a summary of mutations.

### Validation and Benchmarking

We implemented the use of two tools for variant calling, both of which have been reported to have high accuracy for identifying SNVs and indels in germline variants. The results were validated through visual inspection using IGV to remove potential sequencing artifacts.

### Ethical Considerations

The study was conducted in accordance with the ethical standards of the institutional research committee. All participants provided written informed consent before participation in the study.

### Software availability

The workflow, including the configurations and tools, is publicly available on the GitHub repository: https://github.com/digenoma-lab/BRCA.

## Data Availability

All data produced in the present work are contained in the manuscript

https://github.com/digenoma-lab/BRCA

## Author contributions

R.M.S. and A.D.G. contributed to the conceptualization of the study. Methodology was developed by E.V, R.M.S., S.L.H., M.C.M., and A.D.G. Software was developed by E.V., M.M., C.M., and A.D.G. Validation was carried out by E.V. and A.D.G. Formal analysis was performed by E.V., P.J., C.M., and A.D.G. Investigation was conducted by E.V., R.M.S., S.L.H., M.C.M., and A.D.G. Resources were provided by R.M.S., S.L.H., M.C.M.., W.E.D., G.V.S., J.A.R., C.M., and A.D.G. Data curation was handled by E.V., R.M.S., J.F.M., L.J., and A.D.G. The original draft was written by E.V. and A.D.G., while review and editing were contributed by E.V., R.M.S., P.J., W.E.D., G.V.S., J.A.R., J.F.M., L.J., C.M., and A.D.G. Visualization was managed by E.V., P.J., and A.D.G. Supervision, project administration, and funding acquisition were handled by A.D.G.

## Acknowledgments

The supercomputing infrastructure of the High-Performance Computing UOH laboratory (FIC 40059065-0) of the University of O’Higgins and The supercomputing infrastructure of the NLHPC (ECM-02);

