## Supplementary Material for "A workflow for clinical profiling of BRCA genes in Chilean breast cancer patients via targeted sequencing"

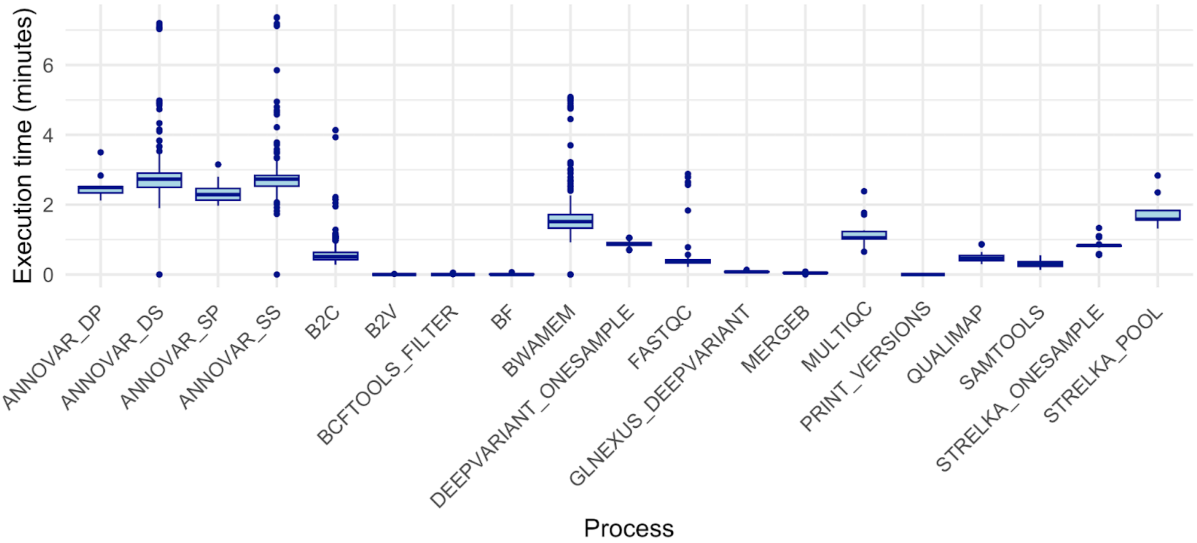


**Figure 1. Execution times of processes in the BRCA Nextflow pipeline.** This visualization shows the distribution of computation times for each process, based on 10 independent executions using 16 sequenced breast cancer samples. The specific processes are as follows: ANNOVAR_DP: Annovar annotation of DeepVariant output in multisample mode; ANNOVAR_DS: Annovar annotation of DeepVariant output in single mode; ANNOVAR_SP: Annovar annotation of Strelka output in multisample mode; ANNOVAR_SS: Annovar annotation of Strelka output in single mode; B2C: BAM to CRAM conversion; BCFTOOLS_FILTER and BCF: VCF filtering; STRELKA_POOL: Variant calling by Strelka in multisample mode; STRELKA_ONESAMPLE: Variant calling by Strelka in single mode; DEEPVARIANT_ONESAMPLE: Variant calling by DeepVariant in single mode; GLNEXUS_DEEPVARIANT: Variant calling by DeepVariant in multisample mode.

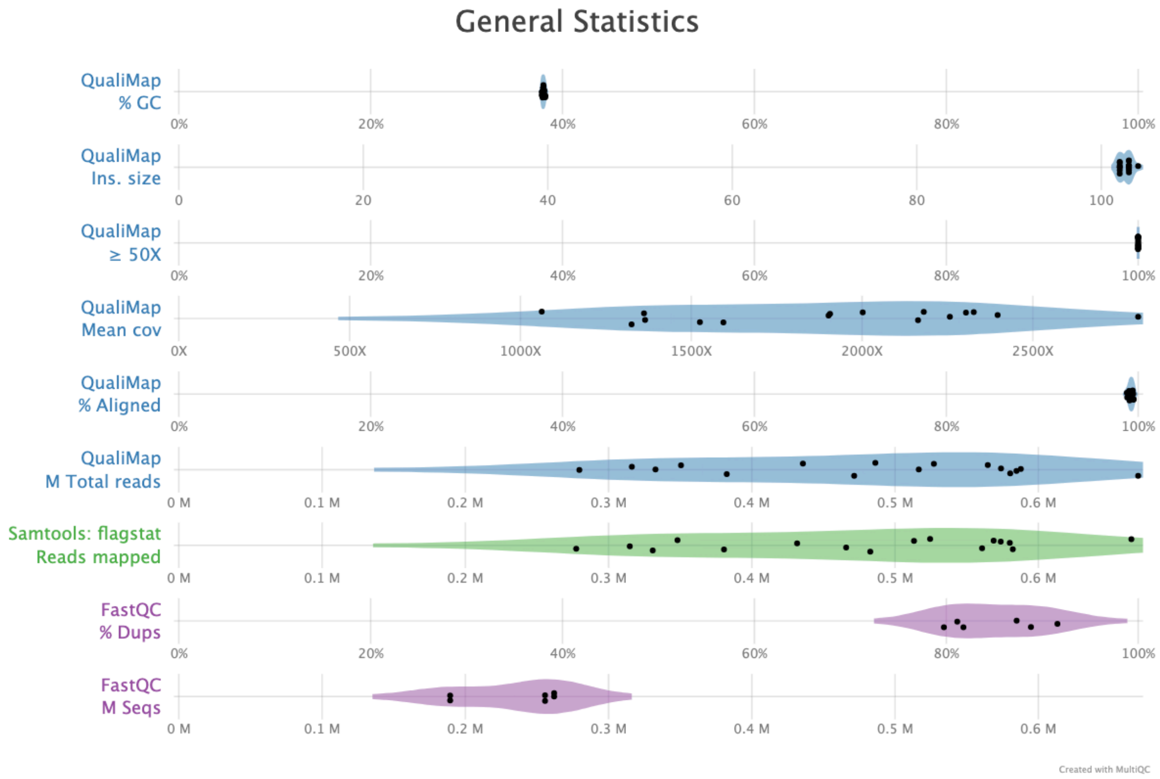


**Figure 2: General statistics of quality metrics for breast cancer patients using the BRCA Workflow in nextflow.** This violin plot, generated by MultiQC tools, summarizes quality metrics across samples obtained from FASTQC, Samtools, and Qualimap. Qualimap provides the following metrics: percentage of CG content, insert size, percentage of reads with at least 50x coverage, mean coverage, and total reads per sample (blue). Samtools indicates the number of mapped reads (green), while FASTQC reports the total number of sequencing reads and duplicated reads (purple). Figure obtained from MultiQC report.

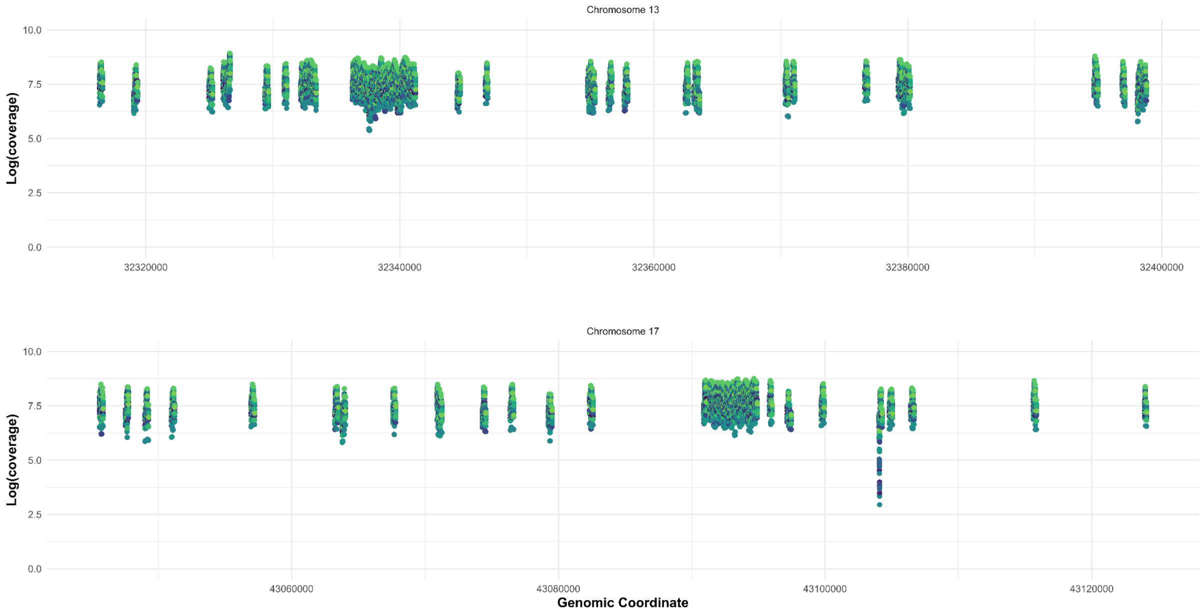


**Figure 3: Coverage by position on target regions of BRCA1/2 genes.** The x-axis represents the genomic coordinates, and the y-axis represents the coverage in logarithmic scale at each on-target position. Each line represents the coverage obtained for patients sequenced. (A) Coverage by position in the *BRCA2* gene. (B) Coverage by position in the *BRCA1* gene.

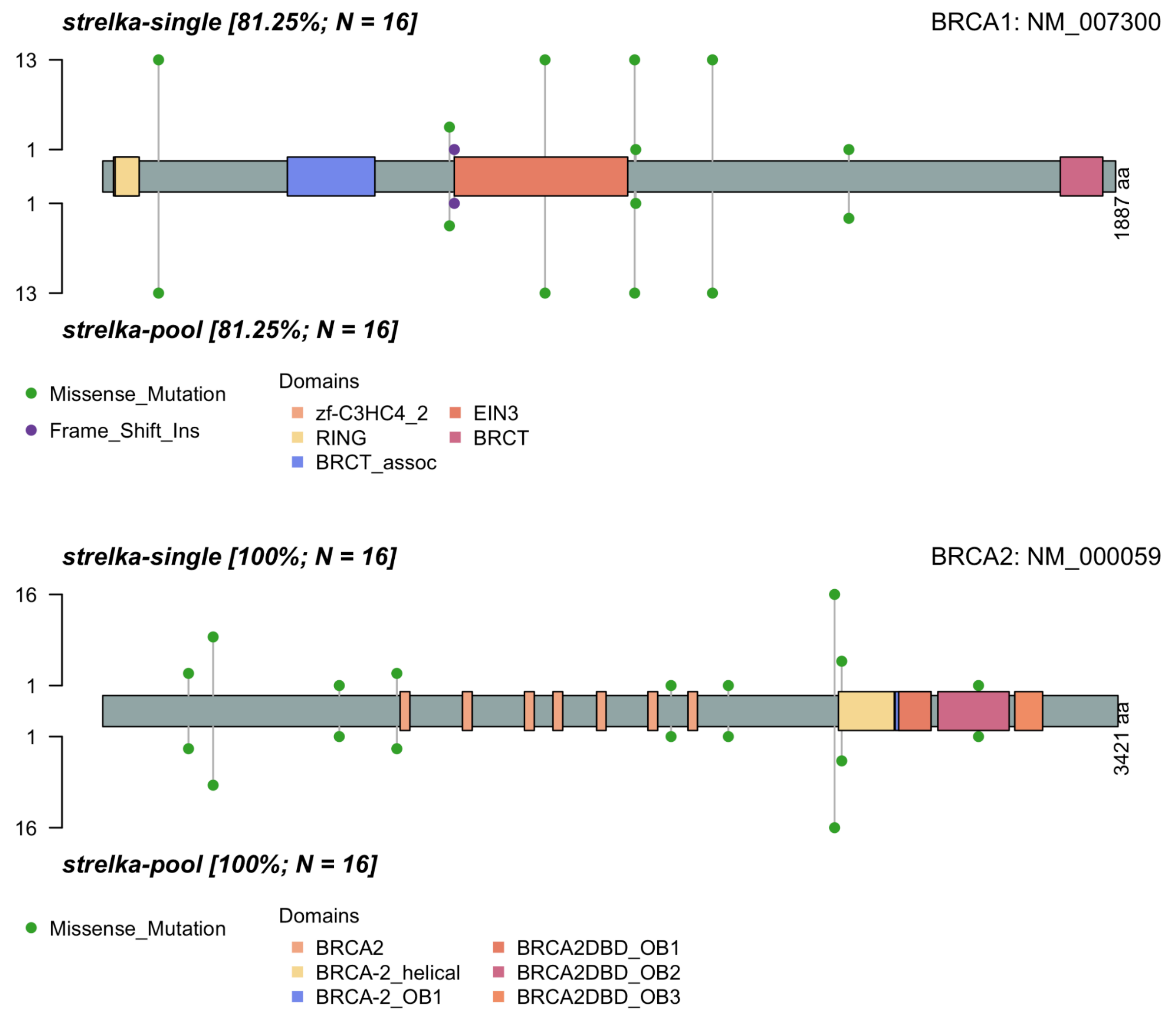


**Figure 4: Comparison of the nonsynonymous variants reported by the Strelka varcaller in single and multisample modes.** SNVs and INDELs reported for the 16 patients in the BRCA1 and BRCA2 genes. The Y-axis shows the number of patients with the mutation, and the X-axis shows the amino acids and domains of each gene.

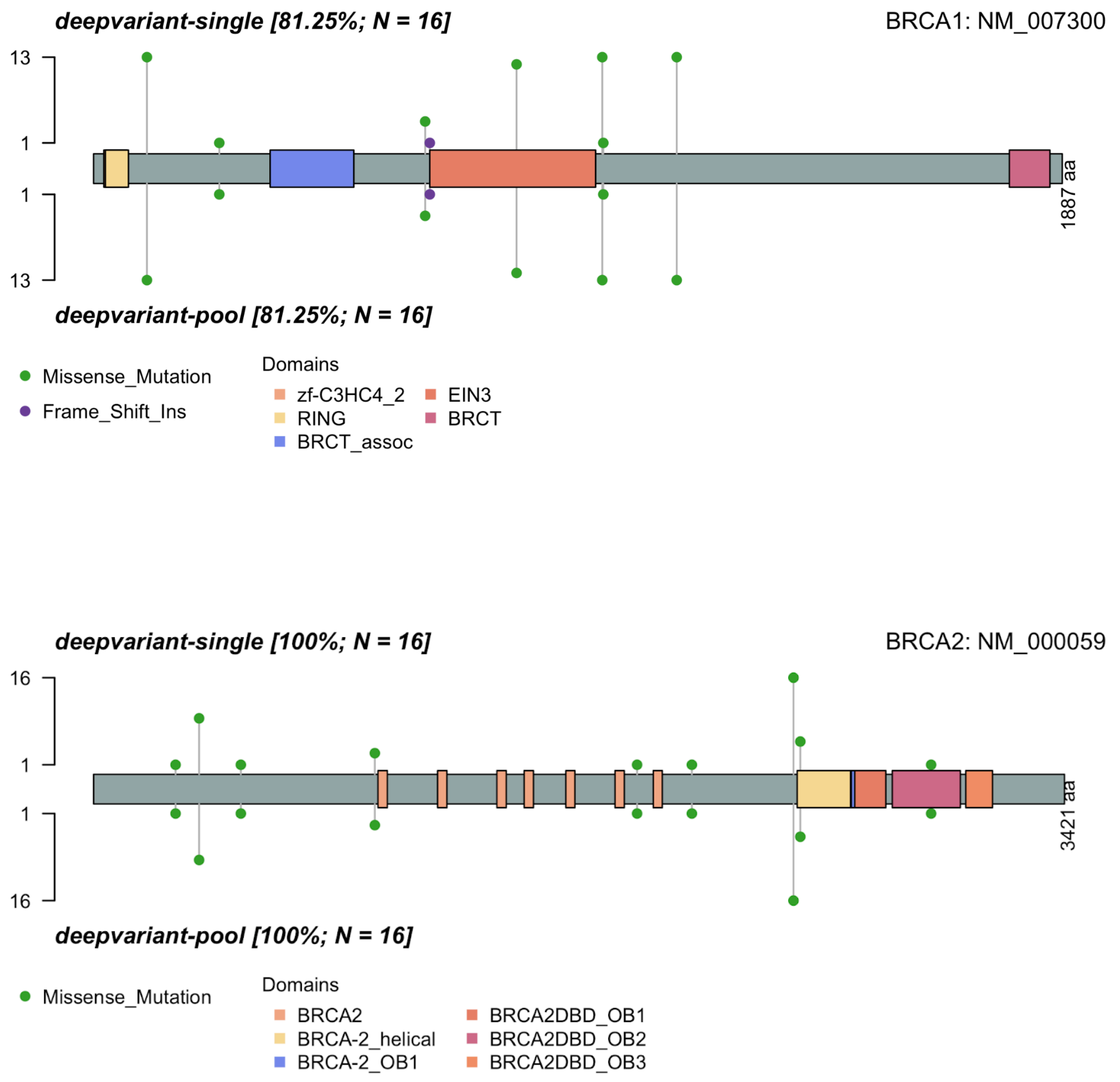


**Figure 5: Comparison of the nonsynonymous variants reported by the DeepVariant varcaller in single and multisample modes.** SNVs and INDELs reported for the 16 patients in the BRCA1 and BRCA2 genes. The Y-axis shows the number of patients with the mutation, and the X-axis shows the amino acids and domains of each gene.

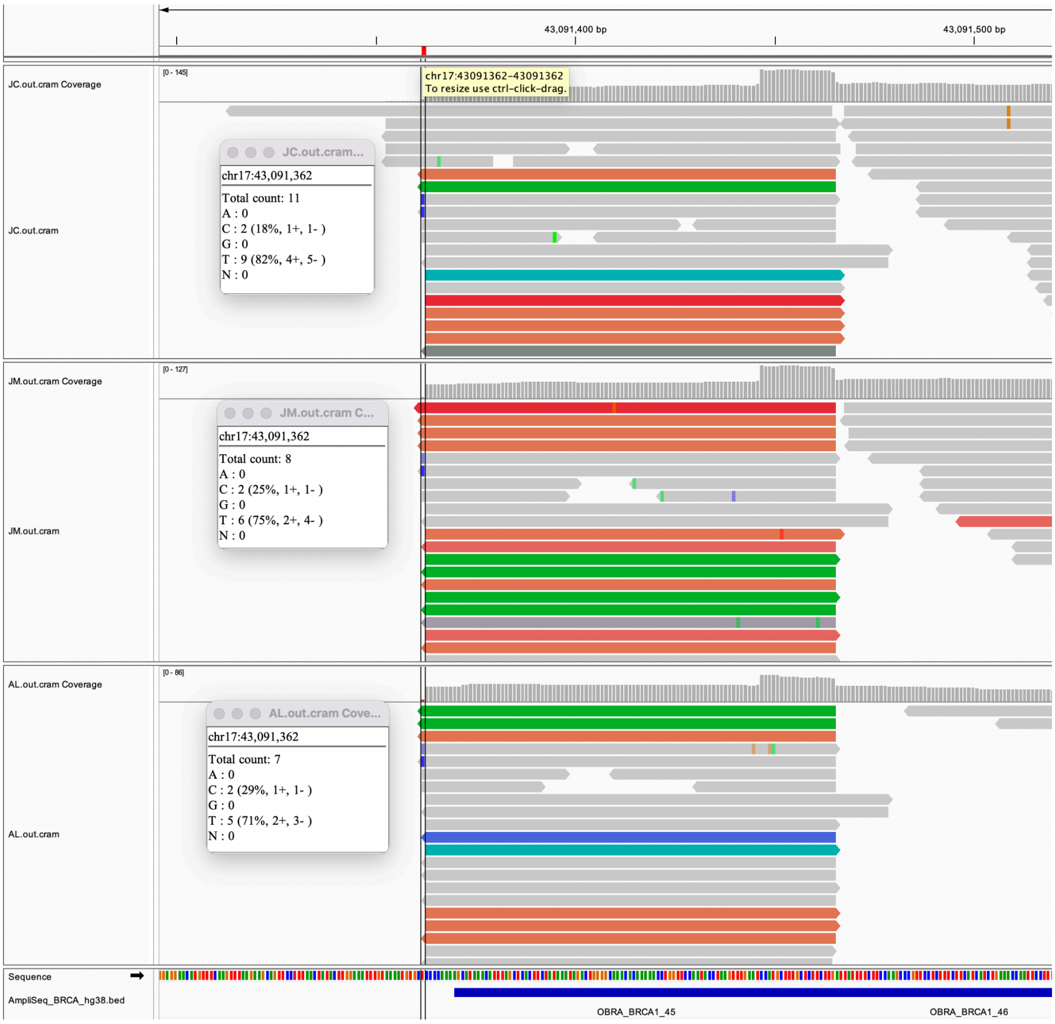


**Figure 6: IGV Visualization of the Missense Mutation E1390G in BRCA1.** Patients JC, JM, and AL reported the novel variant E1390G in *BRCA1*. This variant was only reported by Strelka. Three patients reported this variant in multisample mode, and only JC in single mode. The lower blue line indicates the on-target region of the AmpliSeq BRCA1/2 panel, and the horizontal line indicates the position of the mutation.

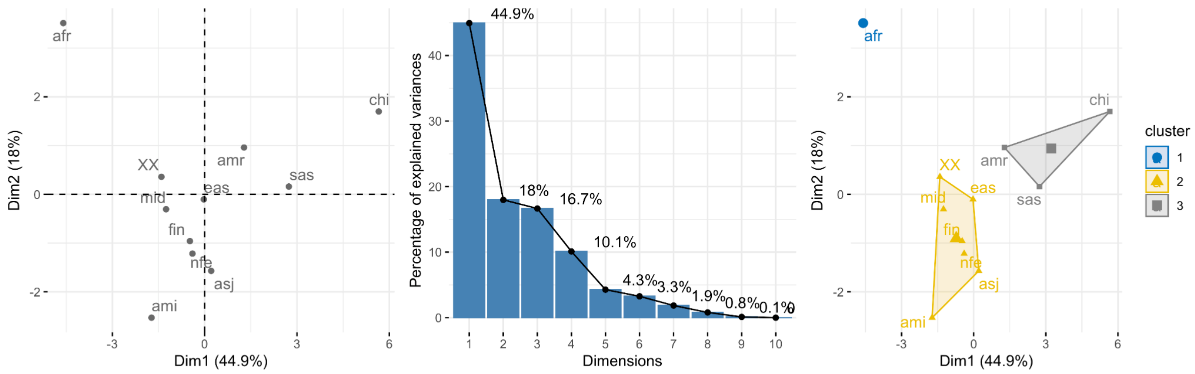


**Figure 7: Principal Component Analysis (PCA) of allele frequency variants in breast cancer patients**. a) PCA plot showing dimensions 1 and 2. b) Percentage of variance explained by each dimension. c) Clustering of populations.
